# Expression of emotion in the British South Asian Diaspora: A Qualitative Study

**DOI:** 10.1101/2022.12.21.22283824

**Authors:** Hira Salman Sharif, Syed K Miah, Amrita Ramanathan, Naomi Glover, Madiha Shaikh

## Abstract

**Background:** ‘Expressed Emotion (EE)’ captures ways in which emotions are expressed within a family environment. Research has found that EE in families has an impact on psychiatric illness, in particular psychosis, such that it increases risk of relapse. EE was conceptualised by research conducted in the UK. Thus, behaviours defined as pathological were largely based on Caucasian samples adhering to UK norms. Cross-cultural variations have been found in the expression of EE and its relationship with clinical outcomes. A more culturally appropriate understanding of norms surrounding the expression of emotion across cultures is required.

**Aims:** This study aims to use a bottom-up approach to provide a culturally specific understanding of family relationships and the expression of emotion across ‘non-clinical’ UK-based South Asian families.

**Methods:** Semi-structured interviews were conducted with 18 South Asian participants to explore their relationships with a significant other. Interviews were analysed using thematic analysis.

**Results:** Four main themes were generated: expression of love, setting boundaries, inter-generational differences and acceptance.

**Conclusion:** The findings indicate considerable cultural variability within emotional expression and highlight the need to interpret EE in the context of socio-cultural norms. Whilst certain expressions of emotion that are considered pathological in Western contexts are present in the UK-based South Asian diaspora, these are perceived as less problematic, indicative of varying cultural norms.

## Introduction

Expressed Emotion (EE) is used to assess the overall quality of the family environment with a focus on social interactions in caregiving relationships (1). Brown (2) characterised five components of EE: critical comments, hostility, emotional over-involvement (EOI), positive remarks and warmth. High EE is defined by increased EOI, hostility and criticism. Research has indicated that higher EE in families impacts patient outcomes across a range of mental health problems (3, 4). In particular, high EE leads to poorer patient mental health outcomes and increases the risk of relapse in psychosis (5).

EE was conceptualised by research conducted in the UK by George Brown, Michael Rutter and colleagues in the late 1950s and 1960s whilst originally trying to determine whether emotions in regular family relationships could be accurately and objectively measured (6, 7). Thus, behaviours defined as pathological were largely based on Caucasian samples adhering to UK norms. Cross-cultural variations have been found in the expression of EE and its relationship with clinical outcomes. Anglo-Americans, for example, display significantly higher EE, including more critical comments and hostility, as compared to their Mexican-American counterparts (8). Moreover, a meta-analysis found that negative outcomes attributed to greater EOI were not observed cross-culturally (9). Rosenfarb et al (10) found that in Black families, higher levels of criticism and intrusiveness were correlated with lower relapse rates. In these families, criticism was seemingly indicative of care and support which consequently decreased patient stress. Another study found UK caregivers of people with schizophrenia and dementia reported greater levels of high EE compared to Japanese caregivers of people with the same conditions (11). Moreover, results showed stark differences in the median number of critical comments across ethnic groups (UK= 6.5 and Japanese = 2). Domain level differences across geographical regions have been highlighted by studies that showed Indian and Chinese samples having higher criticism and warmth compared to Danish and British samples(12, 13). This may be because, unlike in Western cultures, in Chinese cultures, criticism is viewed as a symbol of concern (12).

A recent meta-analysis explored the distribution of EE and its domains across cultures, whilst also assessing the relationship between EE and psychotic relapse(14). Results showed that exposure to high familial EE increased the chance of a relapse by 95% compared to low EE and suggests this relationship is universal. However, there were no significant differences in overall EE scores or domain level scores based on geographical regions. The authors note the categorisation of high and low EE may neglect normative family values and the complexity of culturally defining EE domains. In addition, multiple adjustments to scoring the CFI were made, based on cultural norms, especially for criticism, EOI and warmth. Thus, authors argue against a universal normative EE profile due to the presence of cultural variation in the scoring and interpretation of EE (14).

These findings challenge the universal relationship between high EE and risk of relapse, and thus interventions that aim to reduce EE in families may not always be necessary nor effective across cultures. As such, high levels of certain EE domains may not be detrimental in all cultural contexts. NICE guidelines have recommended using Family Interventions (FI) for psychosis aimed at reducing EE within families, and therefore, decreasing patients’ risk of relapse (15). In the UK, FI is widely used with culturally-diverse populations. Since research has determined that EE varies significantly by culture, both in relation to patient outcomes and prevalence, FI aiming to reduce EE could have adverse effects in non-‘individualistic’ populations (16). This approach may pathologise customary and even protective behaviours in certain cultures (17). A greater understanding of EE and its domains across cultures is therefore necessary to ensure the application of culturally appropriate interventions.

In addition, studies have also found differences in EE rates within the same ethnic groups. For example, an early study comparing high EE rates in families caring for someone with psychosis, found significantly lower rates of high EE in India compared to UK and Denmark (13). Moreover, when the Indian sample was separated into rural and urban inhabitants, high EE rates were considerably lower in rural families (8%) compared to urban (30%). Similar findings were highlighted by another study exploring EE in Chengdhu, China (12), which also noted that people living in the city displayed greater emotional expression than people living in rural areas. Few studies have investigated EE in South Asian populations living in the UK. A study conducted by Hashemi and Cochrane (18) found higher levels of EE in families of British Pakistani (Muslim) patients compared to British Sikh and White families. In both Asian samples, EE did not predict relapse in individuals with psychosis. In a ‘non-clinical’ population (i.e. those without any psychiatric illness), higher levels of EE, especially EOI, were typical in Pakistani Muslims, a characteristic which differed even to Indian Sikhs. Criticism in these families was also significantly higher compared to White families, despite there being no significant differences in criticism in the clinical population. These results show that cultures that appear very outwardly similar may differ in their response to mental illness. They suggest that Pakistani families may generally have intense emotional interactions regardless of whether they are facing obvious stressors; aspects like higher EOI in these families may be indicative of cultural rather than pathological traits. Crucially, the study sample consisted of only Pakistani Muslim and Indian Sikh families; therefore, its application to other South Asian cultures is limited. Moreover, since existing conceptualisations of EE domains (criticism, hostility, EOI) were applied to the South Asian context, it limits the cultural underpinnings of factors that contribute to the quality of a family environment. A more culturally appropriate understanding of norms surrounding the expression of emotion across cultures is required. This study aims to use a bottom-up approach to provide a culturally specific understanding of family relationships and the expression of emotion across ‘non-clinical’ UK-based South Asian families.

## Methods

This study received ethical approval by the Yorkshire and The Humber – South Yorkshire Research Ethics Committee (IRAS: 230098)

### Participants

18 participants were recruited using opportunity and snowball sampling (3 males, 15 females) aged 20 to 39 (M: 28.44; SD: 5.52). UK universities were asked to circulate email invitations to their students and staff, and posts were advertised on social media platforms including Twitter, Facebook and Instagram.

Participants were aged 18 or older, of South Asian ethnicity (nine were of Pakistani ethnicity, six Indian, two Bangladeshi and one Afghan), and residing in the UK. Nine participants were born outside of the UK but had spent 13.4 years on average living in the UK (range=2-31 years) They needed to have a significant other (i.e. child, partner, parent, sibling, etc.) with whom they had regular contact and who they could answer the questions in relation to. Ten participants answered in relation to their parent, five about their partner (husband, fiancé or boyfriend), two about a sibling or sister in law and one about their offspring. If participants or their significant others had any significant mental or physical health problems (i.e. received treatment or diagnosis from a practitioner), they were excluded from the study with the aim of obtaining normative data in the absence of illness-related stressors.

Participants were compensated with £10 Amazon vouchers for their participation.

### Measures

A semi structured interview was conducted by masters level and doctoral researchers. The interview schedule consisted of five open-ended questions exploring participants’ relationships with their ‘significant others’. Existing EE domain terms (eg. emotional over-involvement, criticism) were not included in the questions to allow for an inductive exploration of themes. The schedule was developed based on commonly used tools to identify EE i.e. the Five Minute Speech Sample (19) and Camberwell Family Interview (20).

The interview schedule underwent three iterations which involved discussions amongst the authors to produce the questions and adjust the language used. The interview was piloted with three individuals to determine the order of the questions, whether they were understandable and fit for purpose, and if the interview completion time was appropriate. Adaptations were made to the interview schedule based on the feedback received. A final questionnaire was developed with additional probes which were used to elicit more information where necessary.

### Procedure

Interviews lasted 26 minutes on average (*Min* = 10min 16s; *Max* = 44min 12s), were conducted online via Microsoft Teams, and audio/video recorded. Reflexivity journals were maintained by the interviewers throughout the study process. They also partook in a bracketing interview to engage with the assumptions they were bringing to the process. This was intended to create transparency regarding their position in relation to the research and to remain aware of any potential influences that they may have on the data.

### Data Analysis

Interview transcriptions were attained through Microsoft Teams and proof-read for errors. They were entered into QSR NVivo 7 and analysed using principles of Thematic Analysis (21). This involved six stages: familiarisation, initial coding, searching for themes, reviewing themes, defining, and naming themes and producing the report. Data analysis was conducted using an inductive, bottom-up approach; themes were identified based entirely upon the data collected rather than on pre-existing theoretical concepts.

Generated themes were individually reviewed by the authors in two group discussions before being finalised.

## Results

The data generated four main themes: expression of love, setting boundaries, inter-generational difference, and acceptance. Themes were further categorised into intermediary and sub-themes (see figure 1). Quotations exemplifying each theme are presented in the corresponding tables.

**Figure 1:**
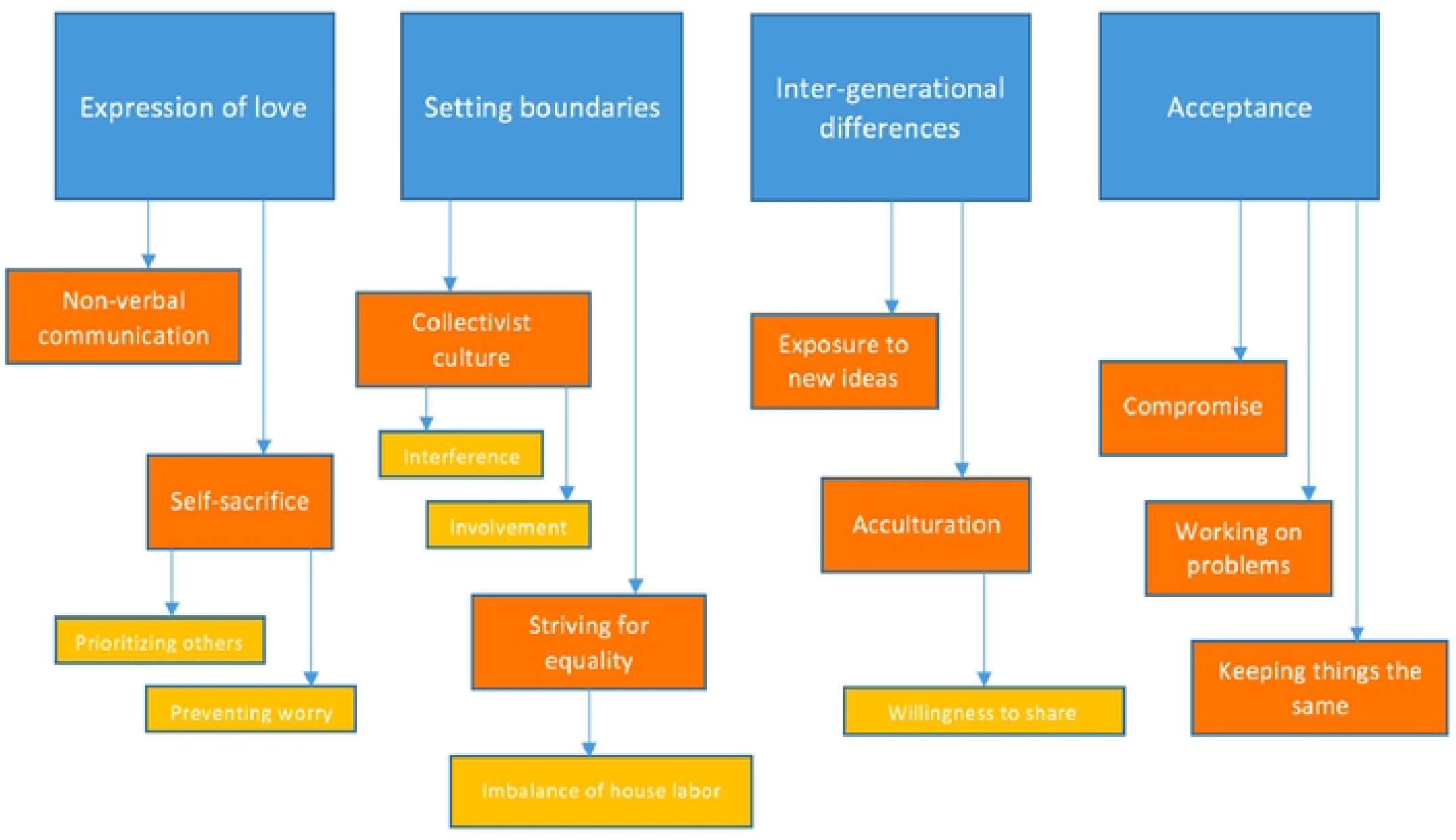
Thematic map (Main themes: Blue; Intermediary themes: Orange; Subthemes: Yellow)

### 1. Expression of love (refer to table 1)

**Table 1.** Quotations illustrating “Expression of love”.

| Main theme | Sub-theme | Quotation |
| --- | --- | --- |
| **Non-verbal communication** |  | And just the fact that she really loves me because, you know, sometimes parents don't always express that, but even though she may not say it specifically, I know that she loves me in the way that she is constantly looking out for me and checking in with me and, just even asking what did I eat today? What did I cook today? It just lets me know that she's thinking of me which is really nice. (P16) |
| **Self-Sacrifice** | Prioritising others | The word selfish is used a lot because it's her way of saying “you're doing things for yourself. You shouldn't. You should just do things for the whole family”. (P11; about sister) |
|  | Preventing worry | I try not to worry her with my worries or with any other things and so even when she's asking “how are you?” I just say everything is good and it's no problem so I try not to share too much and sometimes when I'm holding back, that's difficult as well 'cos, you know, as a daughter, you've always shared your smallest to biggest problem problems with your mother. And I mean we've always been very close and we shared everything with her so it's difficult to hold back and not tell her that this is the problem I'm having these days (P15) |

#### Non-verbal communication

Participants described a lack of overt expression of love within their families. Whilst they were certain ‘love’ was present, they explained how it was rare for it to be communicated explicitly. Instead, love was expressed in non-verbal ways such as constantly looking out for and checking in with your significant others and through spending time and engaging in activities with each other. Participants responded to this lack of verbal expression in one of two ways. Some felt they were lacking something and hoped for it to change. For others, they were content with the unspoken display of affection and felt that the implicit expression of care offered enough security in their relationship, and they were able to experience love and support.

#### Self-sacrifice

Participants described making personal sacrifices for their significant others, owing to a sense of duty which they felt towards their family; this was not specific to any relationship. Whilst acts of self-sacrifice are viewed unfavourably due to their association with poorer mental health outcomes (22), these were a common way of expressing love in South Asian relationships. When individuals did not perform self-sacrificial acts, this was looked down upon.

#### 2. Setting boundaries (refer to table 2)

**Table 2.**
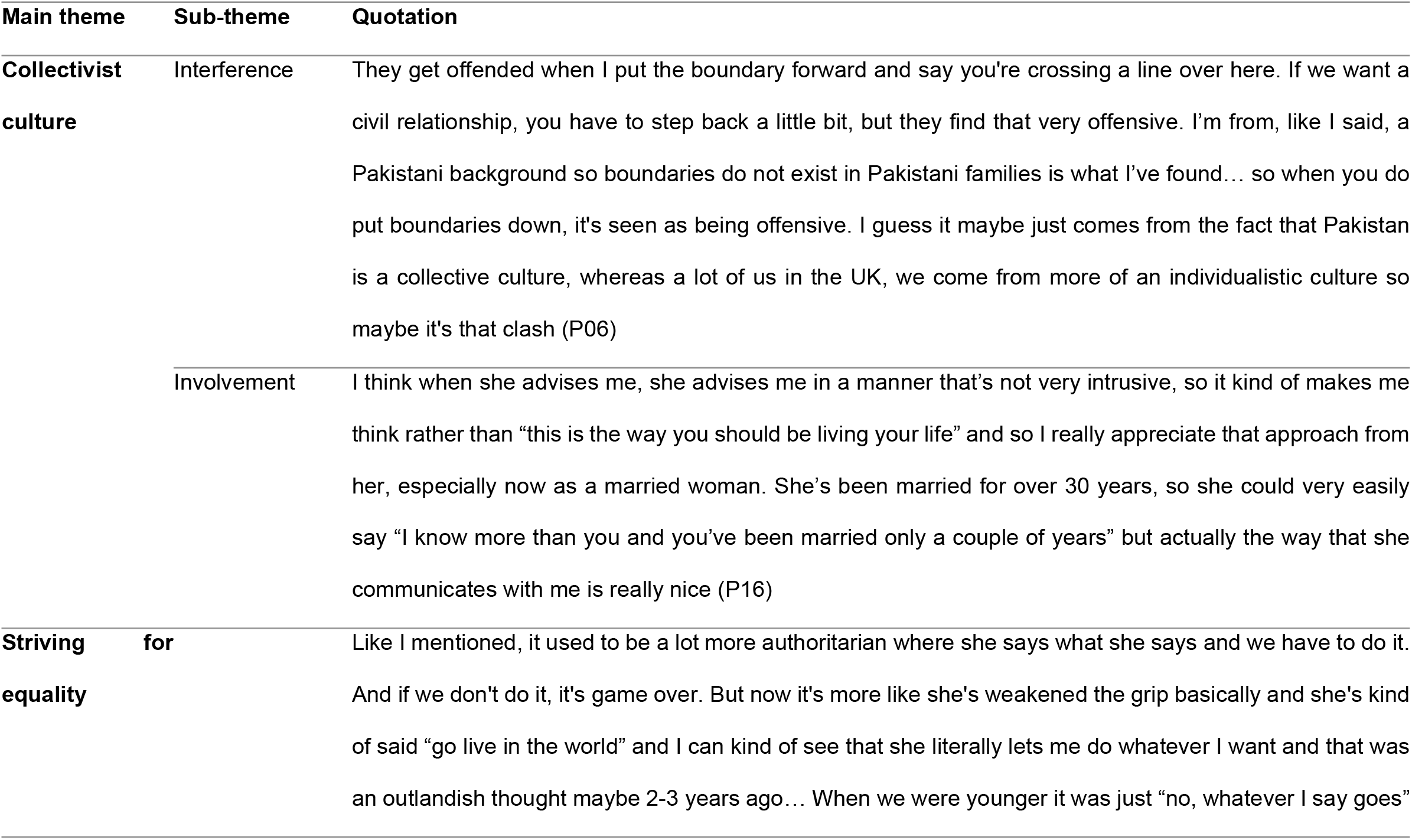
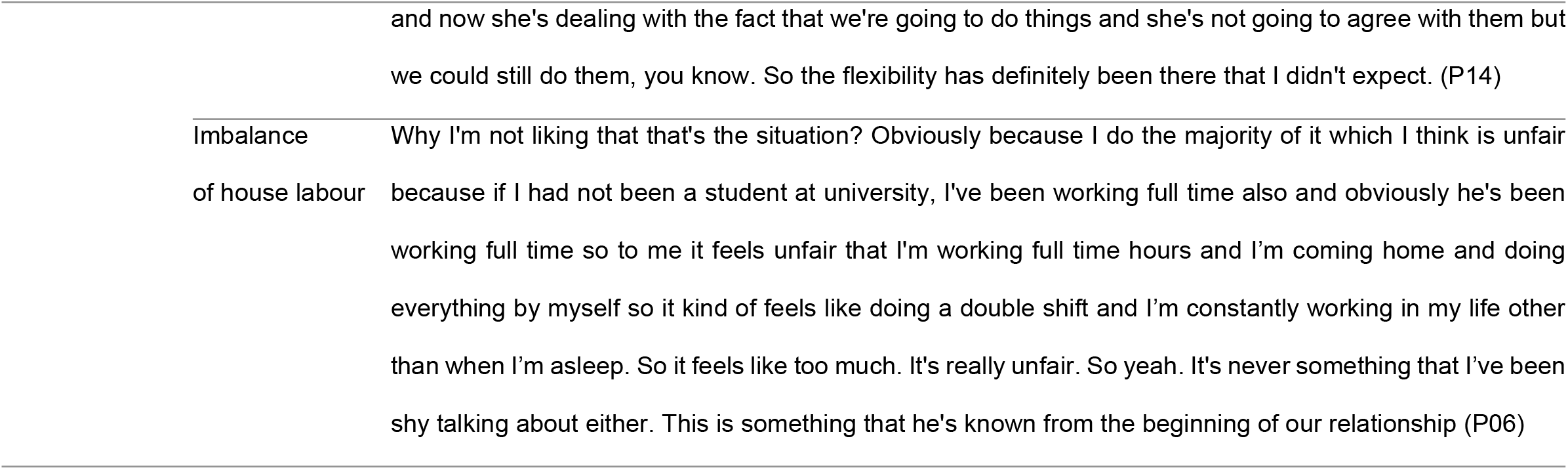
Quotations illustrating “Setting boundaries”.

#### Collectivist culture

Participants described how coming from a collectivist culture meant that families were accustomed to being involved in each other’s lives. Whilst they considered a certain level of involvement acceptable, they did not appreciate *interference* into their daily lives and relationships, possibly highlighting the role of cultural assimilation/acculturation and generational differences.

Problems in relationships primarily emerged when family members were getting overly-involved in their personal lives. This included when individuals did their relatives’ everyday tasks for them, emotionally manipulated them to act in a certain way or offered their opinions on relationships they were not a part of. For instance, *P06* described how her mother-in-law would get involved in her relationship with her husband and took offence when she set boundaries to prevent this. Similarly, *P10* explained how her mother would direct her daughter to act in ways which did not align with her own values. These examples a level of interference or control in these relationships.

Individuals valued *involvement* from their significant others in a friend-like way; positive relationships were “*honest*” and “*informal*”, and those where they could “*banter*” with each other. They appreciated advice from their significant others in a non-intrusive, lenient manner which still granted them autonomy in decision making.

#### Striving for equality

Perhaps due to the collectivist culture, parents stayed involved in their offspring’s lives for longer than they ordinarily would in individualist cultures. Participants described negotiating boundaries with their parents as they got older to make their relationship more adult-to-adult, highlighting the gradual transition from authoritarian parenting to a more equal relationship. The ability to agree-to-disagree provided a good foundation to reduce control and increase flexibility in the relationship.

Participants’ appreciation for the role their parents’ played when they were younger increased with age. As teenagers, they described feeling like their parents were being intrusive or over-protective. As adults, they felt that they could better understand why their parents were like this, suggesting this cohort does value their relatives being involved in their lives.

Women in romantic relationships commonly spoke about the imbalance of house labour as a cause of disagreement, indicative of gender inequality. Especially when both partners were in full-time employment, they felt it was unfair for the household responsibilities to fall on the female partner. They described being open with their partners about their disapproval for this inequality, another way in which boundaries were set.

### 3. Inter-generational differences (refer to table 3)

**Table 3.**
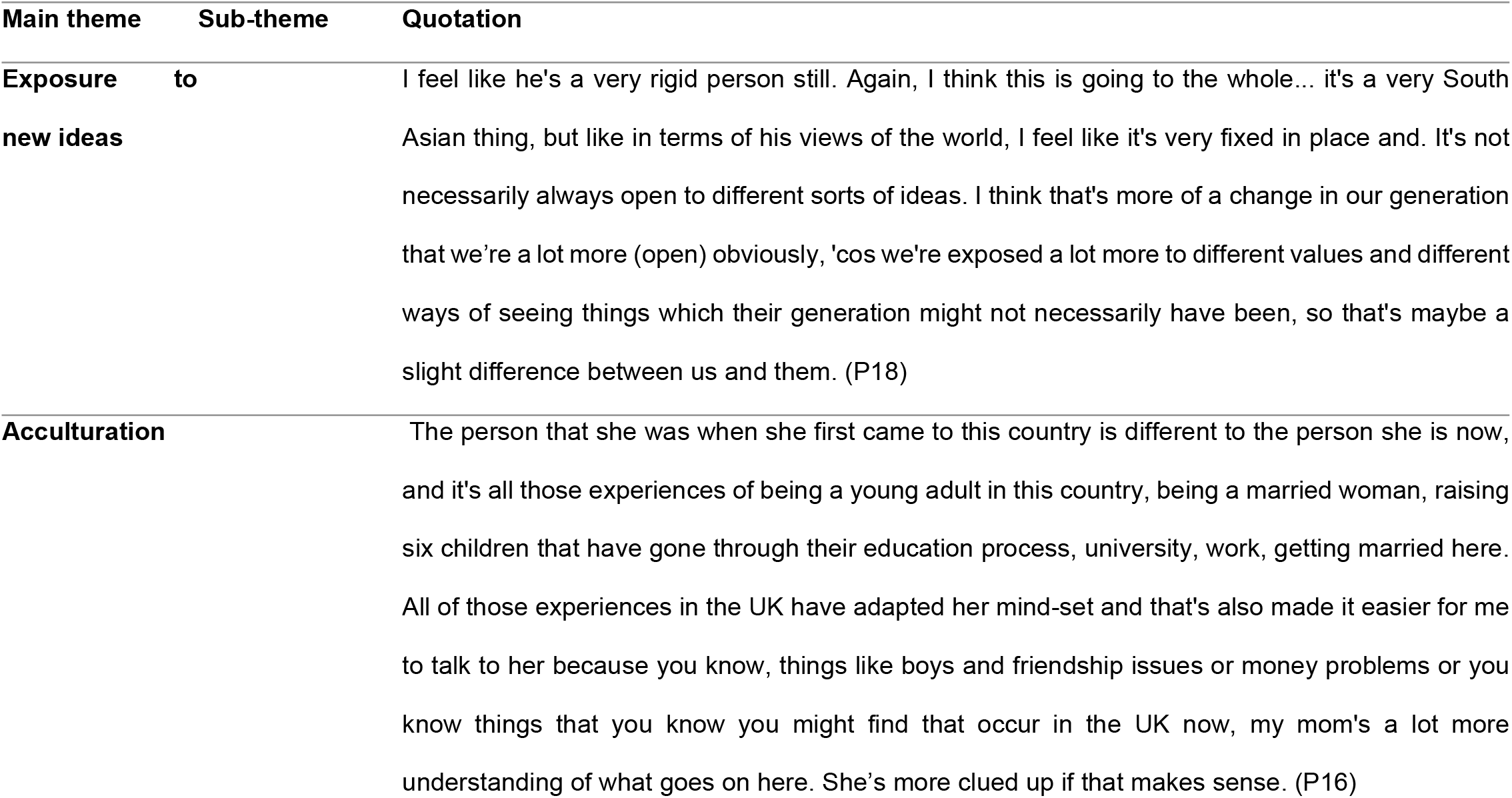
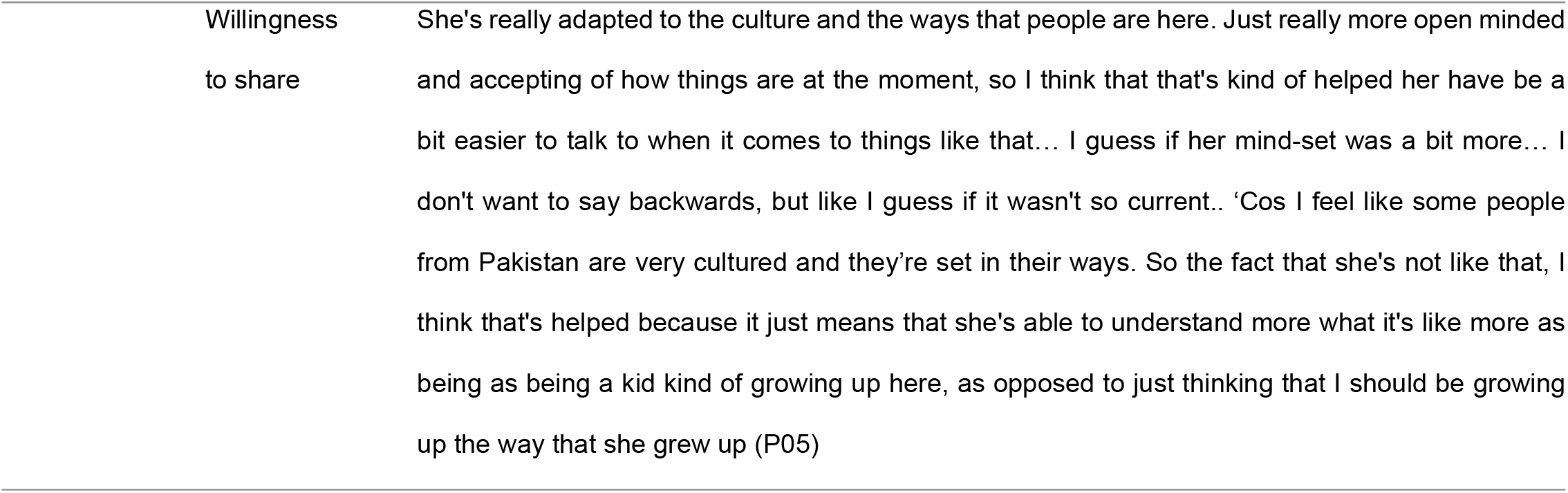
Quotations illustrating “Inter-generational differences”.

#### Exposure to new ideas

Participants explained how elder family members were often quite stubborn in their thinking; they struggled to accept their mistakes or understand other/alternative perspectives. Participants who had grown up in the UK attributed this rigidity to a generational gap. Suggestive of acculturation processes, they felt that they were more open to accepting new ideas as compared to their parents because of the increased exposure they had to other ways of living.

The importance of respect for elders in a South Asian context was also recognised. Regardless of who was factually correct, younger individuals described how they would sometimes have to dismiss their own viewpoints as a sign of respect for their older relatives; these ideas link back to themes of self-sacrifice.

#### Acculturation

A key factor which affected the relationship participants had with their parents was the extent to which they had adapted to Western culture. Having experienced life in the UK, participants described how their parents had become more “*modern*” and “*liberal*” *(P07)*, differing from the general conservative stance most South Asian parents held. It is likely that having lived in both South Asia and the UK, these parents held views aligned with value systems which incorporated both traditional more collectivist and ‘western/individualistic’ concepts. This therefore reduced the extent to which their viewpoints differed from their offspring, and allowed them to be more open about their thoughts and experiences.

Whilst openness was a characteristic which many individuals described as crucial for a healthy relationship, offspring were still mindful about what they shared with their parents. This was especially the case when thoughts or behaviours did not align with their parents’ cultural expectations. There was seemingly more openness in parent-offspring relationships where parents were more willing to tolerate differences in viewpoints and agree-to-disagree.

### 4. Acceptance (refer to table 4)

**Table 4.**
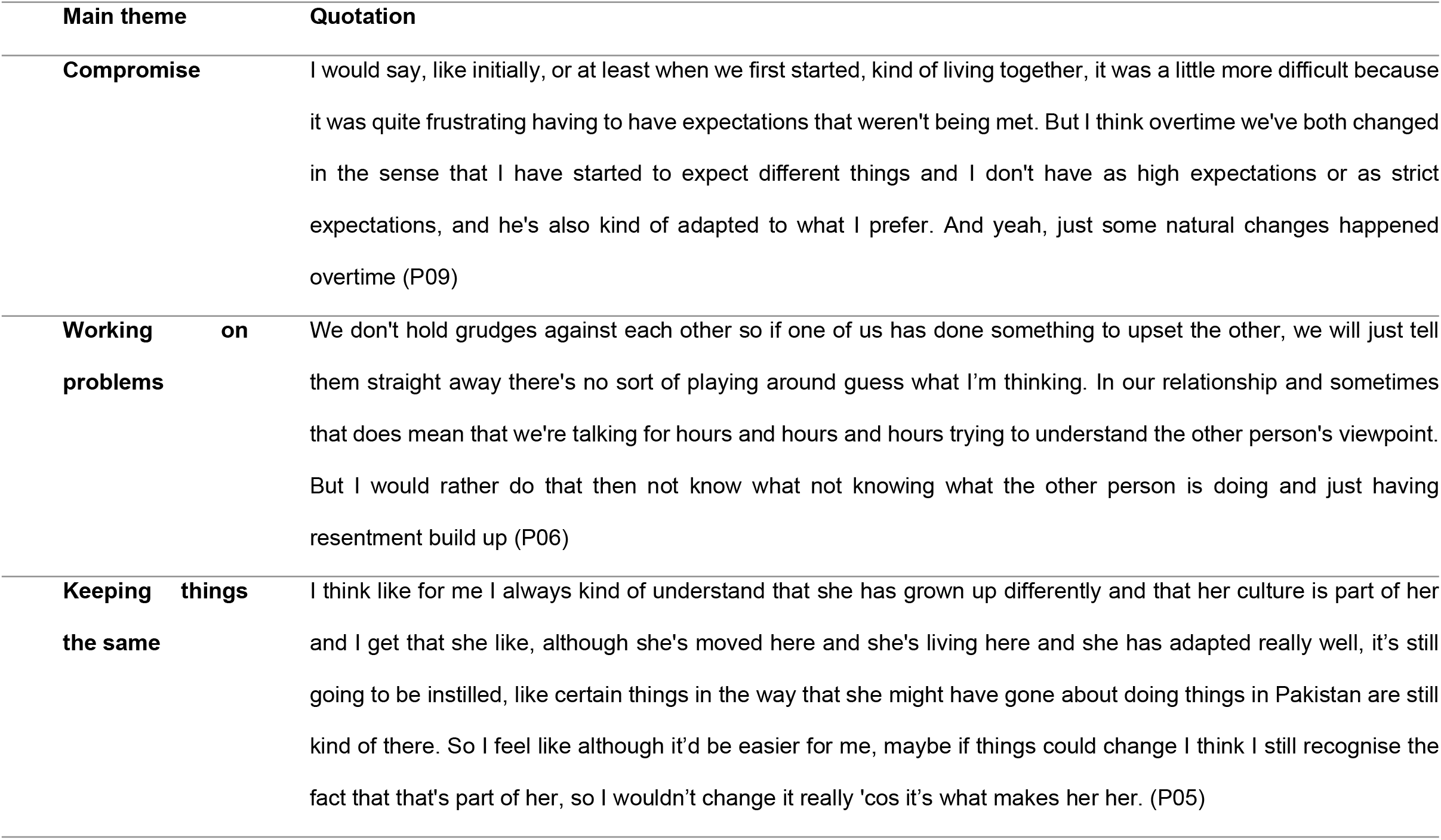
Quotations illustrating “Acceptance”.

| Main theme | Quotation |
| --- | --- |
| **Compromise** | I would say, like initially, or at least when we first started, kind of living together, it was a little more difficult because it was quite frustrating having to have expectations that weren't being met. But I think overtime we've both changed in the sense that I have started to expect different things and I don't have as high expectations or as strict expectations, and he's also kind of adapted to what I prefer. And yeah, just some natural changes happened overtime (P09) |
| **Working on problems** | We don't hold grudges against each other so if one of us has done something to upset the other, we will just tell them straight away there's no sort of playing around guess what I'm thinking. In our relationship and sometimes that does mean that we're talking for hours and hours and hours trying to understand the other person's viewpoint. But I would rather do that then not know what not knowing what the other person is doing and just having resentment build up (P06) |
| **Keeping things the same** | I think like for me I always kind of understand that she has grown up differently and that her culture is part of her and I get that she like, although she's moved here and she's living here and she has adapted really well, it's still going to be instilled, like certain things in the way that she might have gone about doing things in Pakistan are still kind of there. So I feel like although it'd be easier for me, maybe if things could change I think I still recognise the fact that that's part of her, so I wouldn't change it really 'cos it's what makes her her. (P05) |

#### Compromise

Participants discussing romantic relationships commonly recognised that they would have to accept certain details about their partner and their lifestyles to make the relationship work and vice versa. They became more understanding of their partners’ views and actions over time; this helped strengthen their relationship.

#### Working on problems

When problems arose in relationships, participants described how they tried to acknowledge and address them immediately. This was the case in all relationships, romantic and otherwise. Participants valued being able to converse with their significant other about their feelings and needs. They appreciated that their significant others were open to these conversations, and that they could work on making these changes together. Interestingly, this is not the case cross-culturally; other communities value space for reflection before they address a problem (23).

#### Keeping things the same

Interestingly, participants explained how although they disliked certain aspects about their significant other, for instance rigidity or conservative thinking, they did not want these things to change. They worried that changing these traits would affect their relationship.

## Discussion

This study provides an understanding of emotional expression in UK-based South Asian family relationships. It uses an inductive approach to examine how emotions are expressed within this population. These findings provide an awareness of South Asian cultural norms in the UK, in relation to caregiving and receiving behaviours. They contribute to the wider literature on understanding EE through a cultural lens.

Corroborating Hashemi and Cochrane’s (18) report, this study showed that acts of self-sacrifice were common in South Asian families even in the absence of significant health stressors. This raises the question of whether self-sacrificial behaviours in this population are pathological or merely an acceptable way of expressing love. The latter explanation is supported by research suggesting that Pakistani children are raised in a way that encourages closeness and sacrificial behaviours for their relatives (24). In fact, the absence of personal sacrifices and other acts of EOI was perceived as a lack of care (25). Therefore, self-sacrificial acts in South Asian communities may not actually warrant the same level of concern as they would in other socio-cultural contexts where they may be indicative of subjugation and denying one’s own needs to please others (26).

The intermediary theme ‘acculturation’ highlighted how participants’ relationship with their parents was affected by differences in their bicultural identities. When these were similar, participants were more open. Previous literature established an association between generational status (first vs second vs later) and varying acculturation preferences. It found that second-generation Pakistani immigrants favoured an integration strategy compared to their first-generation counterparts who preferred separating from the dominant culture (27). Evidently, these differences impact the expectations individuals have of their relationships. Those who held on to their cultural values had corresponding expectations, such as greater involvement or self-sacrificial behaviours from their significant others. Conversely, those who acculturated more had expectations resembling local values of privacy, autonomy and individualism. This incongruity impacted relationships as individuals had trouble meeting the other’s expectations. Nevertheless, this study found that South Asians held on to certain values (for example, respect for elders) which allowed them to accept, and even appreciate, these differences. The themes of compromise and negotiating and accepting differences are very prevalent in the South Asian diaspora in the UK, especially among the second generation. They reflect the struggles of navigating a bicultural identity and intergenerational differences; these concepts are important to consider when delivering clinical interventions, and to aid communication between family members regarding these disparities.

South Asian families displayed many of the behaviours Brown et al. (28, 29) identified as indicators of warmth. They displayed concern by looking out for and checking up on each other and expressed interest through spending time with their significant others. Only one participant talked about the use of explicit positive remarks in her family. The majority discussed how love was not verbalised in their households, but instead shown in non-verbal ways including involvement, not interference, and self-sacrifice. This corroborates previous research which showed that Pakistani families expressed fewer positive remarks on average compared to those in other communities and highlights cultural variations in the expression of warmth (30).

In collectivist cultures, an absence of positive remarks could have more negative repercussions than the presence of criticism, which might instead be indicative of care and support. Thus tailoring clinical interventions to increase the expression of positive remarks and enhance positive feelings and resilience instead of attempting to reduce criticism might be helpful in this sociocultural context (31).

Participants valued and even expected a degree of involvement from their relatives but distinguished welcomed involvement from interference. They wanted their significant others to take interest in their lives, give them advice and be a source of encouragement. This can be attributed to their collectivist culture (32) where closeness and interdependence are expected in relationships. These communities place family at their core and value their individual needs as secondary to their family duties (33). The distinction between intrusive and non-intrusive involvement is of importance as the former is predictive of psychosis relapse (14). This finding was corroborated by participants in this study who appreciated involvement but not interference.

Only two participants described experiencing criticism and hostility from their significant others. *P11* described “*harsh, mean comments*” from her sister which stemmed from her irritation at her doing more for herself than for the whole family. Similarly, *P03* described passive aggressive comments from her mother about the limited time she spared for her. Both examples demonstrate relationship difficulties stemming from participants placing their own needs before their families. This links to intergenerational differences, highlighting a move away from the expectations of being self-sacrificing and dutiful towards your family to having a more individualistic stance. It also highlights how in the absence of neutral or positive aspects of EE, criticism and hostility can have negative consequences on relationships; a finding which appears to apply cross-culturally (34).

### Limitations

These findings may have been biased by gynocentric views and perceptions. Research has suggested that women display higher emotional over-involvement than males (9) because of the caretaking and attachment role traditionally attributed to them. Although the views of the male participants corresponded with the females, different themes may have emerged in a less gender-biased sample.

The lack of representation across South Asian cultures in the sample could be another limitation that did not allow exploration of within-group variation. Since participants from Nepal, Sri Lanka, Bhutan or the Maldives could not be recruited, it cannot be determined whether their views were represented in our findings. Despite the similarities between South Asian cultures, people in these regions may still differ owing to factors like religion or politics. Even in terms of EE, for instance, studies have shown that Pakistani families had higher EE than Indian families (18).

### Implications and future research

These findings help to distinguish between behaviours that are pathological and customary in UK based South Asian families. For example characteristics such as ‘involvement’ and ‘self-sacrifice’ in South Asian caregiving relationships may serve protective functions. Equally, findings highlight the impact of navigating and negotiating bicultural identities and generational differences in the expression of emotion in the South Asian context.

Future research should apply these findings to explore caregiving relationships in South Asian families that are facing physical or mental health stressors and investigate healthcare professionals’ perspectives and experiences of working with UK-based South Asian families. This can facilitate the development of culturally-specific adaptations to clinical interventions.

## Data Availability

Study materials and data are available upon request.

